# Return to work outcomes in solid organ transplant recipients: a protocol for a global scoping review

**DOI:** 10.1101/2025.02.12.25322136

**Authors:** Vinícius Araújo Pereira, William Donegá Martinez, Martins Fideles dos Santos-Neto, Thaysa Castro Molina, William José Duca, Paulo César Arroyo-Júnior, Allana C. Fortunato, Adília Maria Pires Sciarra, Andressa Karina Stefani, Alex Bertolazzo Quitério, Sônia Maria Maciel Lopes, Luiz Otávio Maciel Lopes, Lucian Borges Lázaro, Flávia Cristina Custódio, Isabela Amaral de Almeida Bistafa, José Nathan Fernandes Rocha, João Daniel De Souza Menezes, Matheus Querino da Silva, Emerson Roberto dos Santos, Licia Paula Schelbauer Borges, Gabriela Gouvea Silva, Vania Del Arco Paschoal, Maysa Alahmar Bianchin, Luís Cesar Fava Spessoto, Heloísa Cristina Caldas, Fernando Nestor Facio Júnior, Júlio César André, Renato Ferreira da Silva

**Affiliations:** FAMERP- Faculty of Medicine of São José do Rio Preto, Brazil; HA - Cancer Hospital of Barretos, Brazil; FUNFARME - São José do Rio Preto Regional Medical School Foundation, Brazil; SCMV - Holy House of Mercy of Votuporanga, Brazil

## Abstract

Solid organ transplantation treats end-stage organ failure, improving longevity and quality of life. Return to work post-transplant is a positive indicator of treatment success. However, labor is influenced by multiple biopsychosocial factors, leading to complex barriers that affect recipients’ opportunities. Mapping return-to-work literature may reveal gaps in conceptualization, instruments, analyses, and key determinants. Following Joanna Briggs Institute methodology, this protocol aims to identify knowledge gaps and map the processes and outcomes in the literature on return to work after liver, kidney, heart, and lung transplantation. Following the Population, Concept, and Context strategy, this review is guided by the research question: “What has the literature shown about return to work after solid organ transplantation?”. This protocol was created and recorded on the Open Science Framework under DOI 10.17605/OSF.IO/Q6HVT. Database selection and search strategy were determined by a librarian specializing in health sciences. The literature search will be conducted in PubMed, Scopus, EMBASE, LILACS, and Web of Science databases. Eligible studies include primary and secondary research, systematic reviews, meta-analyses, clinical trials, case studies, and observational studies on return-to-work post-transplant, in English or Portuguese, with no time restrictions. Two reviewers will independently perform the selection and data extraction. Discrepancies will be resolved through consensus, with a third researcher addressing conflicts. The data will be extracted using a standardized form developed for this review to collect key details about the studies’ origin, context, purpose, content, population, and variables related to the return-to-work process. These data will be synthesized following Synthesis Without Meta Analysis guidelines and summarized narratively using tables, graphs, thematic analysis, and, if feasible, a meta-analysis will be conducted. This scoping review protocol relies solely on publicly available data and does not involve human participants, so institutional review board approval was unnecessary. Ethical aspects of included studies will be assessed in our analysis.

## Introduction

Solid organ transplantation has emerged as the central treatment for organ failure and end-stage diseases involving the liver, kidney, heart, and lung. Its objective is to improve longevity and quality of life while reducing the impact of diseases and the costs associated with care [1]. In this context, in addition to representing an indicator of treatment success, return to work after solid organ transplantation also plays a positive role in financial, psychosocial, and clinical indicators [2-6]. Furthermore, it endorses the importance of investments in vocational rehabilitation programs and the economic sustainability of transplant services [7]. However, the possibility of labor reintegration is conditioned by multiple factors that sometimes involve elements beyond the individual (economic, geographic, political conditions, etc.) [2, 3, 8]. This characterizes challenges and barriers in which labor return is far from uniform for all transplant recipients [1,4].

The assessment and analysis of return to work after transplantation require further validation and the development of standardized instruments, as their absence hinders cross-study comparisons, conceptual clarity, and the identification of key determinants [7]. This is especially relevant as the literature indicates the influence of variables with biopsychosocial scope, such as the effect of immunosuppressants, pre-transplant diagnosis, post-transplant complications, depression, anxiety [1, 4, 6, 7], social security system, access to benefits, age, sex, education, physical effort in professional activities, pre-transplant unemployment, inaccessible labor market, among others [2-4, 6, 9, 10].

A preliminary search in the National Library of Medicine (PubMed), and Scopus (Elsevier) databases was conducted in May 2024, not identifying recent scoping reviews specifically addressing the return to work after multiple solid organ transplants. This scoping review aims to map the existing literature, identify knowledge gaps, and explore key concepts related to return-to-work post-transplantation, providing a foundation for future research, new methods, psychometric instruments, and policies for this crucial area.

The inclusion of these four vital organs in the proposed study’s scope will potentially allow an updated, comparative, and dynamic analysis to promote an understanding of the multifactorial processes and challenges that engender a return to work after transplantation.

Conducting a protocol and a scoping review study investigating the return to work after solid organ transplantation is paramount. In addition to the limited number of recent reviews on this topic, the study set needs to incorporate the joint analysis of the leading solid organ transplanted organs included in the criteria of this protocol. Nevertheless, the mentioned studies also lack critical analyses or possible biases in the scope of return to work [2, 11].

## Materials and methods

### Type of study

This is a scoping review that will be conducted according to the methodological frameworks proposed by the Joanna Briggs Institute (JBI) for scoping reviews [12]. The review process will adhere to the following steps: 1 -formulation of the research question; 2 - identification of relevant studies; 3 - study selection; 4 - data extraction; 5 - synthesis and reporting of results; and 6 - expert consultation.

### Protocol registration

This protocol was created and recorded on the Open Science Framework (OSF) [13] under DOI 10.17605/OSF.IO/Q6HVT.

### Research question

What has the literature shown about return-to-work after solid organ transplantation?

### Objective

To identify knowledge gaps and map biopsychosocial processes and outcomes in the literature on return to work after liver, kidney, heart, and lung transplantation, considering transnational contexts and diverse methodological approaches.

### Inclusion criteria

Studies that centrally analyze the return-to-work process after solid organ transplantation will be included, encompassing primary research such as observational and experimental studies, as well as secondary studies like systematic reviews and meta-analyses. Both qualitative, quantitative, and mixed-methods approaches will be considered. No hierarchy will be established among the types of studies, with all being equally considered for the literature mapping. Eligible studies must be published in English or Portuguese, with no restrictions on publication date or country of origin.

### Exclusion criteria

Studies that contain arbitrary analyses or a limited set of variables on the return-to-work process. Gray literature, including theses and dissertations, as well as duplicate publications, letters to editors, abstracts, and opinion articles, will not be considered. Pediatric and cell transplant studies and any research not specifically related to liver, kidney, heart, or lung transplantation will also be excluded.

### Information sources

#### Research strategy

Widely applied in scoping and systematic reviews, the Population, Concept, and Context strategy was used to formulate the research question [14].

#### Population

The population of interest for this scoping review includes adult (≥18 years) **recipients of solid organ transplants**, specifically liver, kidney, heart, and lung. Studies with participants of all sexes and gender identities will be considered, regardless of pre-transplant employment status. The review will cover studies evaluating return to work at any post-transplant period, from the immediate postoperative phase to the long-term (>5 years). Both participants who were employed prior to transplantation and those seeking employment after the procedure will be included.

#### Concept

The central concept examined in this scoping review is the **return to work after solid organ transplantation**. For the purposes of this study, ‘return to work’ is defined as the resumption of any form of paid employment, whether full-time, part-time, or self-employment, after transplantation. The concept encompasses not only the act of returning to work but also related aspects such as employability, workability, vocational rehabilitation, and job retention post-transplantation.

#### Context

The context of this scoping review is broad and global. **Studies from all geographic regions** will be considered without restrictions, providing a worldwide perspective on the return to work after solid organ transplantation. The review will include research conducted in various settings, such as transplant centers, post-transplant follow-up clinics, workplaces, and communities. Additionally, the review will encompass studies from countries with diverse healthcare systems and transplant policies, enabling comparisons between different models of care and support.

### Databases and search strategy

The search strategies were developed in collaboration with a librarian specializing in health sciences. To ensure precision, accuracy, and comprehensiveness in the research, the controlled vocabulary MeSH was used for the PubMed database and the controlled vocabulary EMTREE for the Excerpta Medica Database (EMBASE) database. The searches will be conducted in May 2024. The strategies may be refined during the review process to optimize the sensitivity and specificity of the search.

The search strategy will follow the three steps recommended by JBI:

1. **Initial limited search:** An initial search will be conducted in the PubMed and Scopus databases to identify relevant articles on return to work after solid organ transplantation. The indexing terms and keywords used in these articles will be analyzed to refine the search strategy.
2. **Expanded second search:** Based on the results of the initial search, a comprehensive search will be carried out across all included databases, using all identified indexing terms and keywords.
3. **Reference list search:** The reference lists of the selected articles will be examined to identify additional relevant studies that may not have been captured through electronic search.

The descriptors used in the search were “Employment,” “Return to Work,” and “Transplantation.” The databases consulted included PubMed, Scopus, EMBASE, Literatura Latino Americana e do Caribe em Ciências da Saúde (LILACS), and Web of Science, ensuring comprehensive coverage of the literature relevant to the research. The search strategy used these descriptors associated with the Boolean operators AND, OR, and NOT, being customized for each database.

The detailed approach to the search strategy is exemplified in Table 1, which outlines the methodology tailored for the PubMed database. Furthermore, this strategy, along with those applied to other databases and the respective number of references retrieved, is available in S1 File. Search Strategy.

**Table 1.**
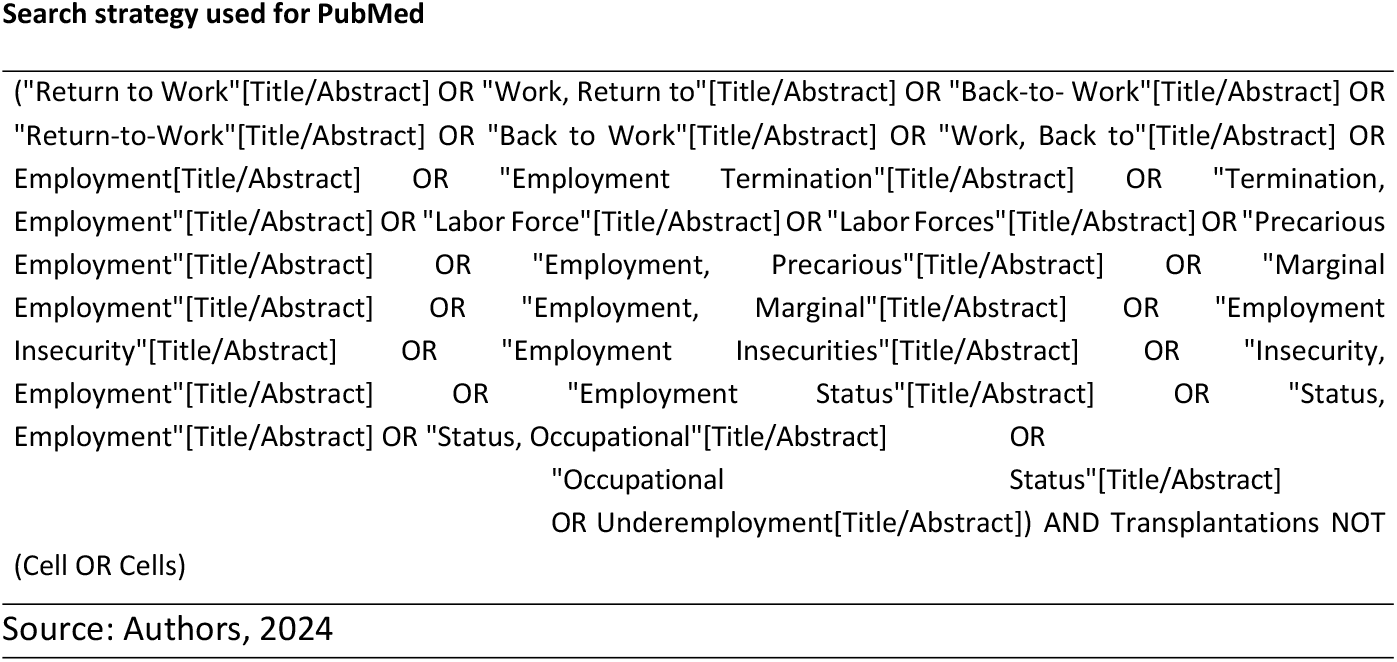
Search strategy in PubMed.

### Study selection

The study selection process will be conducted independently by two reviewers using a double-blinded approach in the Rayyan web tool for reference management [15]. Before starting the selection process itself, the reviewers will participate in a training session to ensure a uniform understanding of the inclusion and exclusion criteria. An initial sample of 50 studies (titles and abstracts) will be reviewed by both reviewers for calibration, and any discrepancies will be discussed to refine the application of the criteria. A standardized selection form will be used to ensure consistency.

Initially, titles and abstracts will be screened, followed by removing duplicates and full-text reading of potentially relevant studies. With the support of the Rayyan platform, the removal of articles that do not meet the inclusion criteria will be recorded and reported in the scoping review. Discrepancies will be resolved by consensus or by a third reviewer. The selection process will follow the recommendations of PRISMA-ScR (PRISMA extension for Scoping Reviews) [16] and will be presented through a flowchart.

### Data extraction

Data will be extracted using a standardized form developed specifically for this review [17]. The form will include information about authorship, year of publication, country, objective, study summary, variables used to characterize the population, variables used to analyze the process of return to work after solid organ transplantation, evaluated outcomes, primary results, and conclusions. These and other variables are available in Table 2.

**Table 2.** Data extraction form.

| Data extraction |
| --- |
| Article title |
| First author |
| Journal |
| Year of publication |
| Country |
| Study type |
| Study objective |
| Methods |
| Sample size (if applicable) |
| Inclusion criteria |
| Exclusion criteria |
| Transplanted organ |
| Living or deceased donor transplant? |
| Main sociodemographic variables (if applicable) |
| Main psychosocial variables (if applicable) |
| Main clinical variables (if applicable) |
| Variables used to measure and analyze return to work |
| Positive factors for return to work |
| Negative factors for return to work |
| Main conclusions |
| Reported biases |
| Additional notes |
| Source: Authors, 2024. |

Data extraction will be conducted independently by two reviewers using a standardized form. Before full extraction, the form will be piloted with a sample of 5 studies to ensure its suitability and make any necessary adjustments. Any discrepancies in the extraction process will be resolved through discussion between the reviewers, with the involvement of a third reviewer if needed. After extraction, a random sample comprising 20% of the studies will have their data verified by a third reviewer to ensure accuracy. Although formal quality assessment of studies is not typical in scoping reviews, notes on the methodological robustness of the studies will be included in the ‘Additional notes’ field of the extraction form.

### Data synthesis and presentation

The data synthesis will be performed in a narrative form, with tables and graphs to summarize the findings [18]. Qualitative analyses, such as thematic analysis, may be performed to identify recurring themes in the included studies [19]. If possible and appropriate, quantitative analyses, such as meta-analyses, will be considered to combine the results of similar studies [20]. To enhance rigor and minimize potential bias, the results will be processed using appropriate software and subjected to statistical analyses. Additionally, peer evaluation will be conducted to identify any potential overlooked biases. The narrative synthesis will follow the guidelines of Synthesis Without Meta-analysis (SWiM) [21], ensuring transparency and reproducibility. Additionally, the IRAMUTEQ software will be used to analyze the textual corpus of the selected articles.

The thematic analysis, if conducted, will follow the six-phase approach proposed by Braun and Clarke [22], using the NVivo software for coding and organizing themes. The heterogeneity among studies will be narratively assessed, considering methodological, population, and contextual differences. Gaps in the literature will be explicitly identified and discussed as potential areas for future research. In case of inconsistencies between studies, we will seek to explain the possible reasons for these divergences, considering factors such as study design, context, and population characteristics.

In addition to the mentioned tables and graphs, we plan to use a conceptual map to illustrate the relationships between the main factors influencing the return to work after solid organ transplantation. A summary table with the main characteristics of the included studies will be presented, highlighting authors, year, country, type of transplant, sample size, and main results. A world map will be used to visualize the geographical distribution of the included studies. Relevant direct quotations from the studies will be incorporated into the narrative synthesis to illustrate key themes. A specific section will be dedicated to presenting the identified gaps in the literature, along with recommendations for future research.

### Consultation

Consultations with experts in the field of solid organ transplantation will be conducted to validate the findings and provide additional insights [1]. A panel of 5-7 experts, including transplant surgeons, occupational rehabilitation specialists, and representatives of transplant patient associations, will be selected based on their experience and relevant publications. The experts will be recruited through email invitations, and their feedback will be integrated into the final synthesis of the results, contributing to the interpretation and contextualization of the findings.

The consultation process will be conducted after the initial data synthesis, comprising individual semi-structured interviews and a subsequent online focus group session to validate findings and refine interpretations. The interviews and group sessions will be recorded, transcribed, and thematically analyzed. The feedback from the experts will be documented in a summary table, indicating how each suggestion was incorporated into the final synthesis or, if not incorporated, the justification for such a decision.

This scoping review will strictly follow the PRISMA-ScR (PRISMA extension for Scoping Reviews) guidelines to ensure transparent and comprehensive reporting [16]. The PRISMA-ScR checklist will be used as a guide throughout the review process and the preparation of the final report. All 22 items of the checklist will be addressed, with particular emphasis on elements related to the review rationale, search strategy, study selection process, data extraction, and result synthesis. The completed PRISMA-ScR checklist will be included as an appendix in the final report, allowing readers to easily assess the review’s compliance with these guidelines. No significant adaptations to the PRISMA-ScR guidelines were made for this review, thus ensuring full adherence to the established standard.

### Study timeline

The scoping review is proceeding according to the following schedule:

1. Protocol Development and Registration (completed: October 2023).
2. Search Strategy Refinement (completed: February 2025):
  • The search strategy was finalized in collaboration with a health sciences librarian.
3. Initial Database Searching (in progress: March-April 2025):
  • Systematic searches are being conducted across multiple databases.
4. Title and Abstract Screening (anticipated: May-June 2025):
  • Two independent reviewers will assess titles and abstracts using the Rayyan platform.
5. Full-Text Review and Data Extraction (anticipated: July-August 2025):
  • Full-text articles meeting the inclusion criteria will be retrieved and reviewed.
  • Data extraction will be carried out using a standardized pre-developed form.
6. Data Synthesis and Analysis (anticipated: September-October 2025):
  • Thematic analysis and data synthesis will be performed.
7. Manuscript Preparation and Submission (anticipated: November-December 2025).

The anticipated results, expected by December 2025, may include a comprehensive mapping of the literature on return to work after solid organ transplantation, identification of gaps in existing knowledge, and recommendations for future research and instrument development.

## Data Availability

No datasets were generated or analysed during the current study. All relevant data from this study will be made available upon study completion.

https://doi.org/10.17605/OSF.IO/Q6HVT

## Ethics statement

This study is a scoping review protocol that does not involve direct human participant’s research. It utilizes only publicly available, previously published information. Therefore, institutional review board approval was not required or sought for this study. The ethical considerations of the studies included in our review will be critically assessed and reported as part of our analysis.

## Supporting information

**S1 File. Search Strategy**.

(DOCX)

